# COVID-19 among bartenders and waiters before and after pub lockdown

**DOI:** 10.1101/2021.02.01.21250905

**Authors:** Fredrik Methi, Kjetil Telle, Karin Magnusson

**Affiliations:** Norwegian Institute of Public Health, Cluster for Health Services Research, Oslo, Norway; Lund University, Faculty of Medicine, Department of Clinical Sciences Lund, Orthopaedics, Clinical Epidemiology Unit, Lund, Sweden

## Abstract

**Aim:** To study how different bans on serving alcohol in Norwegian bars and restaurants were related to the detection of SARS-CoV-2 in bartenders and waiters.

**Methods:** In 24,276 bartenders and waiters and 1,287,970 persons with other occupations (mean [SD] age 41.7 [12.8] years and 51.7% men), we examined the weekly rates of workers tested and detected with SARS-CoV-2, one to five weeks before and one to five weeks after implementation of different degrees of bans on serving alcohol in pubs and restaurants, across 56 Norwegian municipalities with: 1) full blanket ban, 2) partial ban with hourly restrictions (e.g. from 10 pm), or 3) no ban, adjusted for age, sex and testing behavior.

**Results:** In municipalities introducing *full ban*, COVID-19 among bartenders and waiters had been reduced by 65% by three weeks (from 3.4 [95%CI=2.5-4.3] to 1.2 [95%CI=0.7-1.7] per 1000), i.e. to the same levels as that for persons with other occupations (1.8 [95%CI=1.7-1.9] vs 1.2 [95%CI=1.1-1.3] per 1000). Similarly, in municipalities introducing *partial ban*, COVID-19 among bartenders and waiters had been reduced by 68% by three weeks (from 2.5 [95%CI=1.4-3.6] to 0.8 [95%CI=0.0-1.5] per 1000). However, there was more uncertainty to the estimated reduction for partial bans.

**Conclusion:** Municipalities with higher levels of confirmed COVID-19 among bartenders and waiters implemented stricter bans on serving of alcohol than other municipalities. Contraction of COVID-19 among bartenders and waiters declined similarly in municipalities with full and partial bans.

## Introduction

The emergence of the coronavirus disease 2019 (COVID-19) in late 2019 and 2020 has led to an extraordinary social and economic upheaval across the globe. The SARS-Cov-2 virus has been assumed to spread easily in pubs and restaurants, because of lack of social distancing, consumption of alcohol, and load talk, which may increase the spread of respiratory droplets [1-2]. In sum, these factors may have put bartenders and waiters at a much higher risk of contracting the virus than persons in other occupations.

We recently reported that bartenders and waiters are among the occupations with the highest COVID-19 incidence in Norway [3]. In March 2020, a large number of COVID-19 cases across Europe could be traced to après-ski bars in Ischgl in Austria [4]. Clusters of virus outbreaks have also been related to pubs and restaurants in Japan and Thailand [1-2]. Thus, strict limitations on citizens’ behavior have been enforced in most countries to mitigate the spread of the virus, including partial or full bans on the serving of alcohol in pubs and restaurants. Despite its widespread use, the effect of different restrictions imposed on bars, pubs and restaurants in reducing the spread of the virus is unknown. It is also unknown *how strict* restrictions should be in order to reduce the spread of the virus, i.e. whether full bans on serving alcohol are required to limit the spread of SARS-CoV-2 among bartenders and waiters, or whether partial bans are enough.

Improved knowledge of the effect of social distancing measures, such as limitations on the serving of alcohol, may be important for stopping the spread of the virus in both bartenders and waiters, and the general population. It might also be important knowledge to owners of restaurants and bars, to protect their workers.

Using longitudinal population-wide data from Norway, we aimed to study associations between bans on serving of alcohol in Norwegian bars and restaurants, and the detection of SARS-CoV-2 in bartenders and waiters when compared to workers in other occupations.

## Methods

### Data sources and population

As part of the legally mandated responsibilities of The Norwegian Institute of Public Health (NIPH) during epidemics, an emergency preparedness register (BeredtC19) was established in cooperation with the Norwegian Directorate of Health to provide rapid knowledge of the epidemic. We utilized individual-level data from BeredtC19, which contains information from the Norwegian Surveillance System for Communicable Diseases (MSIS), The Norwegian Population Registry, the Employer- and Employee-register, the administrative record systems of all Norwegian hospitals (NPR), etc. We merged data using an encrypted version of the unique personal identification number provided all Norwegian residents at birth or upon immigration, to identify detected SARS-CoV-2 across occupations and municipality of residence. The data sources are updated daily, except for the Employer-Employee-register, which was updated on August 25^th^ 2020. Institutional board review was conducted, and the Ethics Committee of South-East Norway confirmed (June 4th 2020, #153204) that external ethical board review was not required.

### Occupation: Bartenders and waiters vs. other occupations

Bartenders and waiters were identified as persons having at least one of the International Standard Classification of Occupation (ISCO-08) codes 5131 (waiters) or 5132 (bartenders) in the Employer- and Employee-register, as registered in week 34, 2020 (August 17^th^– August 23^rd^). In the comparison group, we included all other Norwegian residents between 20 and 70 years with at least one registered other occupation in the same week.

### Outcome: COVID-19

The outcome variable in this study, COVID-19, was identified as either having a confirmed positive polymerase chain reaction (PCR) test and/or by having ICD-10 diagnostic code U07.1 of confirmed COVID-19 in a hospital record.

### Definition of blanket ban on serving alcohol

Data on national and local restrictions that were valid during fall 2020 were gathered from national authorities, newspapers and the respective municipalities, and classified by the time they went into effect, as well as their level of strictness [5-7]. The period from September 2020 to December 2020 was selected because of data availability and because the local variation in bans were the greatest during this period. These data were classified as follows:

1. National ban: There was a national ban on serving alcohol after midnight (12 pm) in place between August 8^th^ (week 32) 2020 and October 12^th^ (week 42) 2020, and between November 7^th^ (week 45) 2020 – January 4^th^ (week 1) 2021 (ordinary restrictions in between these two periods, typically disallowing serving of alcohol after 3 am).
2. Local ban: We classified local variations of restrictions in three groups, using information from 56 municipalities: (1) full ban (i.e. no serving of alcohol allowed), (2) partial ban (e.g. no serving of alcohol after 10 pm), and (3) no ban (i.e. only national restrictions as described above). We included all 52 municipalities in the two most populous counties in Norway (Viken and Oslo), and also the municipalities Bergen, Stavanger, Trondheim and Kristiansand to capture all ten most populous municipalities in Norway. Of the 56 municipalities, 28 imposed local restrictions at various times in 2020. Bergen imposed their restrictions in week 45; Oslo and 17 other municipalities in week 46; four municipalities in week 49; Øvre Eiker in week 51; and four municipalities in week 52 (E-Table 1).

### Statistical analyses

To examine the association between the alcohol serving restrictions and detection of COVID-19 in bartenders and waiters, we calculated the weekly rates of testing and detection of SARS-CoV-2 before and after imposing restrictions in each municipality. We treated the dependent variable, detection of SARS-CoV-2 as a binary variable (yes/no) denoting whether each individual was positive in the respective week.

First, we estimated the weekly rate with its 95% confidence interval [CI] of contracting the virus for bartenders and waiters vs. for persons in other occupations for all 56 municipalities, i.e. our entire population, from calendar week 39 through 52 of 2020. Results from this analysis may inform on overall trends and associations with national restrictions. Second, we disaggregated the data to examine the associations between local restrictions and COVID-19. Municipalities implemented their full or partial ban on serving alcohol at different calendar weeks. Also, some municipalities did not implement any ban. For these municipalities, we randomly assigned hypothetical implementation weeks to enable them to serve as comparison group (see E-Table 1). In this way, we could transform all the calendar weeks to relative weeks for each municipality (including those with no ban) by setting the week the restriction in question was implemented in the municipality to be the relative week 0. For each level of restriction (full, partial and no ban), we then estimated rates of COVID-19 (95% CI) from 5 weeks prior to week 0, to 5 weeks after week 0, for bartenders and waiters vs. other occupations. Available testing is a requirement for detecting SARS-CoV-2, and we performed logistic regression analyses adjusted for testing behavior (testing negative for COVID-19 in the week in question (yes/no)) as well as age and sex. Standard errors were clustered on the individual. Finally, we repeated all analyses stratified on bartenders and waiters in separate analyses. The statistical software used was STATA MP v.16.

## Results

Of an overall 2,562,240 persons aged 20-70 years living in Norway on January 1^st^ 2020 who were employed by week 34, 2020, we studied the 1,312,246 persons living in the 56 municipalities in populous regions of Norway. Their mean [SD] age was 41.7 [12.8] years and 51.7% were men. Of these, 6,209 were employed as bartenders and 18,067 as waiters.

Between week 39 and 52, 2020, 13,875 cases of COVID-19 were notified among the employees, of whom 463 were bartenders (N=128) or waiters (N=335). In the same period 532,028 negative tests were reported in our study population, with 10,465 being from bartenders (N=3,071) and waiters (7,394).

### Overall patterns

Results from the analysis including all waiters, bartenders and other occupations in all the 56 municipalities showed that the rate with COVID-19 was higher among bartenders and waiters than among persons with other occupations, across the time period (Figure 1). The steepest increase for bartenders and waiters was seen in week 45 to 46. Additional measures were imposed both nationally and locally in large parts of Norway in week 45 and 46 [5-7], and we observed a decline in infection, especially among bartenders and waiters, after these weeks (Figure 1).

**Figure 1:**
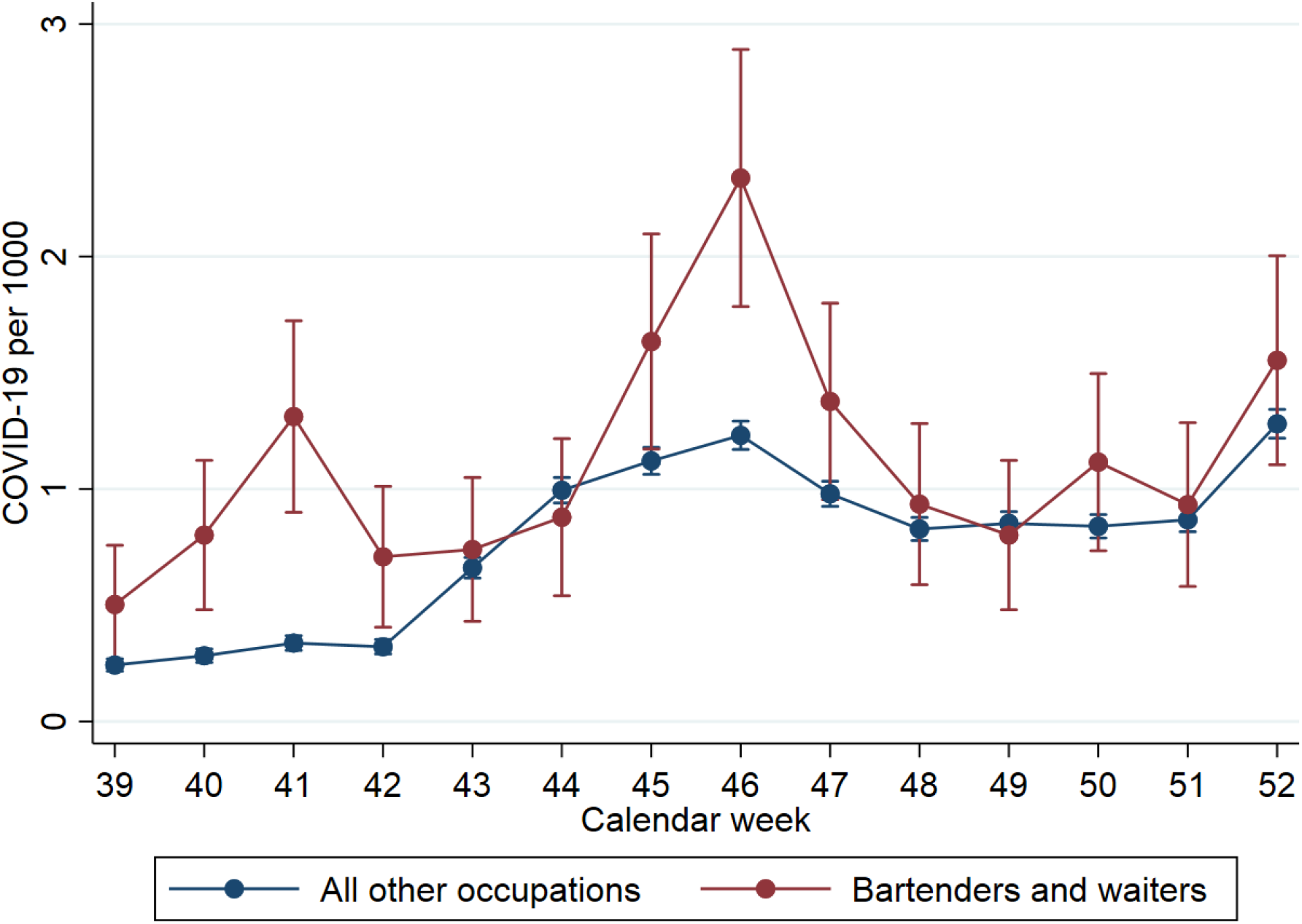
Estimated rate (95% confidence intervals) with confirmed COVID-19 per 1000 in 56 municipalities in Norway for bartenders and waiters (red line) and all other occupations (blue line) by calendar week, adjusted for age, sex and testing.

### Local variations

Of the 56 municipalities, 12 imposed a full ban (covering 667,108 persons) and 16 a partial ban (covering 389,290 persons) on serving of alcohol, at various times. The remaining 28 municipalities did not impose any local restrictions (covering 255,848 persons) (Figure 2). Oslo and surrounding areas imposed the strictest restrictions, i.e. they had a full blanket ban on serving alcohol (Figure 2, E-Table 1).

**Figure 2:**
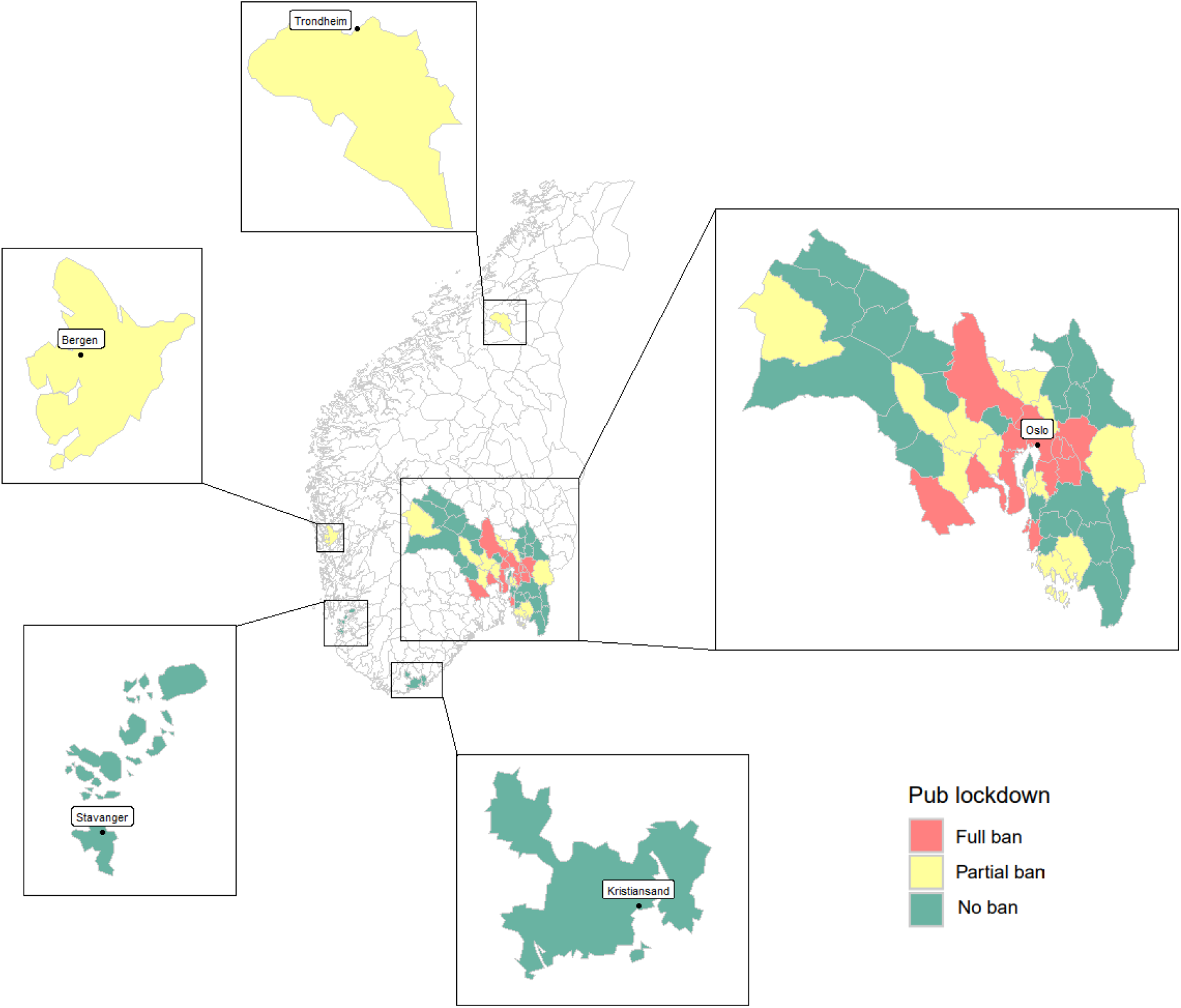
Map of all the 56 municipalities included in the study, with color codes showing the type of ban on serving alcohol. Red colored municipalities imposed a complete ban on serving alcohol. Yellow colored municipalities imposed partial restrictions (e.g. ban on serving alcohol after 10 pm). Green colored municipalities did not impose local restrictions (i.e. they followed only the compulsory national restrictions).

When weeks were relabeled to relative time from the week the restrictions were implemented, we could compare the rates of COVID-19 across municipalities with full versus partial vs no alcohol serving bans (Figure 3a-c). In municipalities that implemented a full ban (Figure 3a), the rate with COVID-19 for bartenders and waiters (3.4 per 1000, 95% CI=2.5-4.3) was higher than that for persons in other occupations (1.8 per 1000, 95% CI=1.7-1.9) in the week of implementation, declining by 65% (to 1.2 per 1000, 95% CI=0.7-1.7) by three weeks after the implementation to the same levels as for persons in other occupations (1.2 per 1000, 95%=CI 1.1-1.3).

**Figure 3.**
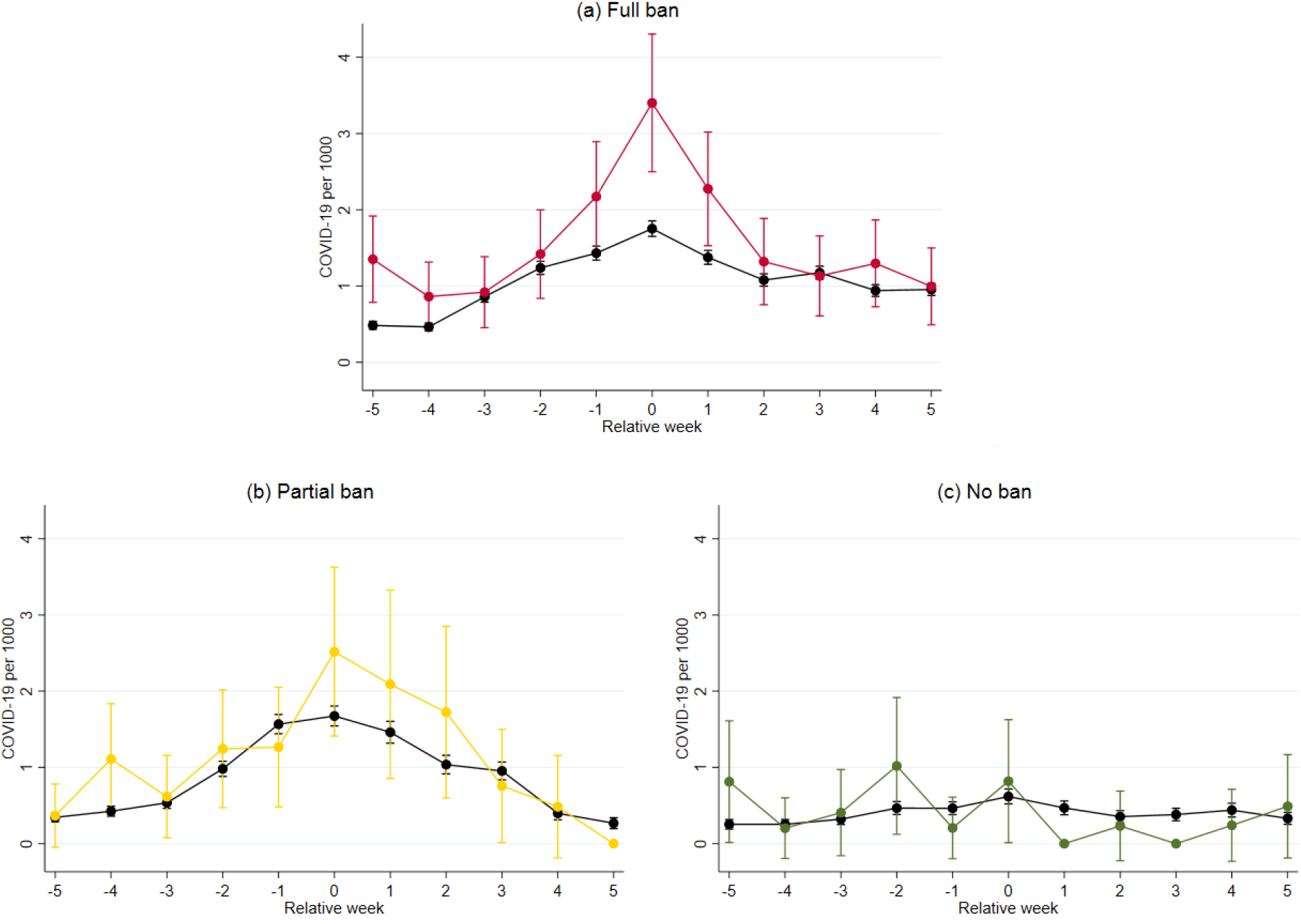
Estimated rates of confirmed COVID-19 per 1000 for bartenders and waiters (colored lines) and all other occupations (black lines) per week, from 5 weeks before to 5 weeks after the week when the municipality of residence implemented partial or full ban on serving of alcohol in bars and restaurants. For municipalities with no restrictions implemented, a hypothetical week of implementation was set by random draw (see Methods). Figure 3(a) shows the rate in municipalities with a full blanket ban on serving alcohol. Figure 3(b) shows the rate in municipalities with partial restrictions (e.g. ban on serving alcohol after 10 pm). Figure 3(c) shows the rate in municipalities with no local restrictions. Estimates based on logistic regression analyses adjusted for age, sex and testing.

In municipalities that implemented partial bans (Figure 3b), the rate with COVID-19 for bartenders and waiters (2.5 per 1000, 95% CI=1.4-3.6) was higher than that for persons in other occupations (1.7 per 1000, 95% CI=1.5-1.8) in the week of implementation. COVID-19 among bartenders and waiters had been reduced by 68% (to 0.8 per 1000, 95% CI=0.0-1.5) by three weeks, to the same levels as for persons in other occupations (1.0 per 1000, 95% CI=0.8-1.1). In municipalities with no restrictions and a randomly assigned hypothetical implementation week (Figure 3c), the rate for bartenders and waiters (0.8 per 1000, 95% CI=0.0-1.6) was variable yet similar to that of persons in other occupations (0.6 per 1000, 95% CI=0.5-0.7) both in the implementation week and three weeks after the hypothetical implementation (0.0 per 1000, 95% CI=0.0-0.0 for bartenders and waiters, and 0.4 per 1000 95% CI=0.3-0.5 for other occupations). Similar results were observed in separate analyses of bartenders and waiters, however, there was more uncertainty to these estimates (E-Table 2).

## Discussion

In this study of 1,312,246 persons including all bartenders, waiters and other workers in 56 municipalities in the most populous regions of Norway, we report that partial and full bans on serving alcohol are associated with a lower incidence of COVID-19 among bartenders and waiters. We also find evidence that partial bans, such as closing pubs, bars and restaurants at 10 pm, are similarly associated with declining rates of detected COVID-19 among bartenders and waiters as full bans. These findings may be of importance for informing local and national authorities in their implementation of restrictions, as well as for companies in the catering and night life business to help protect their workers and customers.

To our knowledge, the current study is the first to assess effects of full or partial bans on serving alcohol during the COVID-19 pandemic. By using routinely collected population-based data and applying longitudinal panel data methods, we may have limited bias due to attrition, selection, confounding and misclassification - strengthening the transparency of causal inference from our study. As an example, data on occupation was available for everyone, and we could adjust for potentially different testing and infection patterns for COVID-19 among bartenders and waiters vs persons in other occupations. We found no previous studies for comparison of the potential effects of bans on serving alcohol. However, our findings confirm our previous reports showing that COVID-19 transmission is disproportionally higher among bartenders and waiters [3]. Our findings also shed new light on previous studies, as we show that transmission is reduced once hourly or full restrictions on serving alcohol are implemented, adding to the evidence that physical distancing interventions are associated with a reduced incidence of COVID-19 [8].

Our findings may be important for pandemic policy. We found indications that hourly restrictions, or partial blanket bans on serving alcohol from 10 pm may reduce the incidence of COVID-19 among bartenders and waiters (Figure 3b). However, there was more uncertainty to the estimates from these analyses of partial bans than there was for analyses of full ban, in which we observed a more certain reduction (Figure 3a). Along the reduction among bartenders and waiters after the two types of alcohol bans, there was also a reduction in COVID-19 among persons in other occupations (Figure 3a, 3b). This reduction in rates of COVID-19 could not be observed in municipalities with no implemented ban, neither among bartenders or waiters, nor among persons in other occupation (Figure 3c). Altogether, these findings may have two different explanations: Either, that full and partial bans on serving alcohol are effective in reducing transmission among persons with *any* occupation (i.e. both bartenders and waiters *and* other occupations), or that measures only are implemented at the top of a transmission curve, which would decline regardless of the implemented measures.

Along this line, some important limitations should be mentioned. First, we could not rule out that infection rates might have declined regardless of the imposed restrictions. While this decline might in theory be related to the restrictions in serving of alcohol, it may also be a sign that the restriction was implemented when the local infection rate was peaking. As an example, although the analysis shows that a partial ban may be similarly associated with declining infection rates as a full ban, infection rates were higher in areas imposing a full ban on serving alcohol, and we cannot say whether implementing a partial ban in these areas would have had the same effect. Fully randomized designs would be required to exclude this potential bias.

Second, restrictions on alcohol serving were rarely or never imposed alone. As an example, in the same week as the nationwide restrictions on alcohol were imposed (week 45), the government also encouraged people to limit social contact and avoid unnecessary domestic travels. In the following week, a constrain of maximum 50 persons in indoor events without fixed seating and 200 with fixed seating, was effectuated [6]. It is likely that also the municipalities with partial vs. full bans implemented such co-measures, whereas the municipalities with a hypothetical implementation week did not. Although the larger decline for bartenders and waiters than for other occupations links our findings to bars and restaurants, there is no way to ensure that our results do not reflect changes in the infection rates that are related to all these measures combined. Relatedly, we capture the municipality of residence of the employees, and some may not be working in the same municipality as they live, for whom the combination of restrictions they have faced are additionally involved.

In conclusion, we report that partial bans on serving alcohol in bars and restaurants may be similarly associated with declines in confirmed COVID-19 among bartenders and waiters as full bans. Considering the burden of full bans to owners and workers in bars and restaurants, hourly restrictions of serving alcohol, i.e. from 10 pm, may be explored further in reducing the spread of the SARS-CoV-2.

## Supporting information

ICMJE form Methi

ICMJE form Magnusson

ICMJE form Telle

## Data Availability

Data are not publicly available.

## Acknowledgements

We would like to thank the Norwegian Directorate of Health, in particular Director for Health Registries Olav Isak Sjøflot and his department, for excellent cooperation in establishing the emergency preparedness register. We would also like to thank Gutorm Høgåsen, Ragnhild Tønnessen and Anja Elsrud Schou Lindman for their invaluable efforts in the work on the register. Our acknowledgements also to Atle Fretheim for critically evaluating our manuscript. The interpretation and reporting of the data are the sole responsibility of the authors, and no endorsement by the register is intended or should be inferred. We would also like to thank everyone at the Norwegian Institute of Public Health who has been part of the outbreak investigation and response team.

## Author contribution

Fredrik Methi had access to all of the data in the study and takes full responsibility for the integrity of the data and the accuracy of the data analysis. Fredrik Methi and Kjetil Telle performed the statistical analyses and Karin Magnusson drafted the manuscript. All authors contributed with acquisition of data, conceptual design, analyses and interpretation of results. All authors contributed in drafting the article or critically revising it for important intellectual content. All authors gave final approval for the version to be submitted.

## References

[1] Kripattanapong S, Jitpeera C, Wongsanuphat S, Issarasongkhram M, Mungaomklang A, & Suphanchaimat R. Clusters of Coronavirus Disease (COVID-19) in Pubs, Bars and Nightclubs in Bangkok, 2020. OSIR Journal. 2020;13(4):146–153

[2] Furuse, Y., Sando, E., Tsuchiya, N., Miyahara, R., Yasuda, I., Ko, Y., et al. Clusters of coronavirus disease in communities, Japan, January–April 2020. Emerging infectious diseases, 2020;26(9):2176–9

[3] Magnusson K, Nygård K, Methi F, Vold L, Telle K. Occupational risk of COVID-19 in the 1st vs 2nd wave of infection. medRxiv2020.10.29.20220426v2 [Preprint]. 2020. Available from: https://doi.org/10.1101/2020.10.29.20220426

[4] Correa-Martínez CL, Kampmeier S, Kümpers P, Schwierzeck V, Hennies M, Hafezi W, et al. A pandemic in times of global tourism: superspreading and exportation of COVID-19 cases from a ski area in Austria. Journal of clinical microbiology. 2020;58(6):1–3.

[5] Verdens Gang. CORONA-VIRUSET: Nasjonale og lokale tiltak i Norge [Internet]. [Updated 28 Jan 2021; cited 28 Jan 2021]. Available from: https://www.vg.no/spesial/corona/tiltak/

[6] Regjeringen. -Hold dere hjemme, ha minst mulig sosial kontakt [Internet]. [Updated 5 Nov 2020; cited 13 Jan 2021]. Available from: https://www.regjeringen.no/no/aktuelt/-hold-dere-hjemme-ha-minst-mulig-sosial-kontakt/id2783763/

[7] Oslo kommune. 9. november: Byrådet har vedtatt sosial nedstenging av Oslo [Internet]. [Updated 11 Nov 2020; cited 13 Jan 2021]. Available from: https://www.oslo.kommune.no/politikk/byradet/pressemeldinger/9-november-byradet-har-vedtatt-sosial-nedstenging-av-oslo

[8] Islam N, Sharp SJ, Chowell G, Shabnam S, Kawachi I, Lacey B, et al. Physical distancing interventions and incidence of coronavirus disease 2019: natural experiment in 149 countries. BMJ 2020;370:m2743.

