## Supplementary tables and figures for "COVID-19 among bartenders and waiters before and after pub lockdown"

By Methi et al., 2021


| | |
|---|---|
| Supplementary Table A: Overview of local restrictions | p. 2-3 |
| Supplementary Figure A: Estimated rates of confirmed COVID-19 for bartenders | p. 4 |
| Supplementary Figure B: Estimated rates of confirmed COVID-19 for waiters | p. 4 |

**Supplementary Table A:** Overview of local restrictions by municipality, type of restriction (1 = no local restrictions; 2 = partial ban; 3 = full ban) and week of implementation. Municipalities with no ban (1) was randomly assigned a hypothetical week of implementation (in parentheses) to allow us to use them as a comparison group.

| Municipality | Restriction type | Week |
|---|---|---|
| Aremark | 1 | (46) |
| Asker | 3 | 46 |
| Aurskog-Høland | 2 | 46 |
| Bergen | 2 | 45 |
| Bærum | 3 | 46 |
| Drammen | 3 | 46 |
| Eidsvoll | 1 | (46) |
| Enebakk | 3 | 46 |
| Flesberg | 1 | (46) |
| Flå | 1 | (49) |
| Fredrikstad | 2 | 49 |
| Frogn | 2 | 46 |
| Gjerdrum | 1 | (46) |
| Gol | 1 | (46) |
| Halden | 1 | (46) |
| Hemsedal | 1 | (52) |
| Hol | 2 | 52 |
| Hole | 1 | (46) |
| Hurdal | 1 | (46) |
| Hvaler | 2 | 49 |
| Indre Østfold | 1 | (46) |
| Jevnaker[1] | 2 | 46 |
| Kongsberg | 3 | 52 |
| Kristiansand | 1 | (46) |
| Krødsherad | 1 | (46) |
| Lier | 2 | 46 |
| Lillestrøm | 3 | 46 |
| Lunner | 2 | 46 |
| Lørenskog | 3 | 46 |
| Marker | 1 | (45) |
| Modum | 2 | 46 |
| Moss | 3 | 49 |
| Nannestad | 1 | (49) |
| Nes | 1 | (46) |
| Nesbyen | 1 | (49) |
| Nesodden | 1 | (52) |
| Nittedal | 2 | 46 |
| Nordre Follo[2] | 3 | 46 |
| Nore og Uvdal | 1 | (49) |

| | | |
|---|---|---|
| Oslo | 3 | 46 |
| Rakkestad | 1 | (46) |
| Ringerike | 3 | 52 |
| Rollag | 1 | (52) |
| Rælingen | 3 | 46 |
| Råde | 1 | (46) |
| Sarpsborg | 2 | 49 |
| Sigdal[3] | 2 | 46 |
| Skiptvet | 1 | (51) |
| Stavanger | 1 | (46) |
| Trondheim | 2 | 52 |
| Ullensaker | 1 | (52) |
| Vestby | 1 | (46) |
| Våler | 1 | (46) |
| Øvre Eiker | 2 | 51 |
| Ål | 1 | (46) |
| Ås | 2 | 46 |

Note: The random assignment was conducted so that the share of municipalities with ban ( 2 and 3) within each implementation weeks was similar to the share of municipalities without ban (1) within the same (actual) implementation weeks. Thus, municipalities without ban are only attributed to hypothetical weeks that are also actual implementation weeks for the municipalities with ban.

[1]Jevnaker removed their partial ban on alcohol in week 50.

[2]Nordre Follo changed from a full ban to a partial ban in week 51

[3]Sigdal removed their partial ban on alcohol in week 52

**Supplementary Figure A:** Estimated rates of confirmed COVID-19 for bartenders separately (colored lines) and all other occupation (black lines) per week.

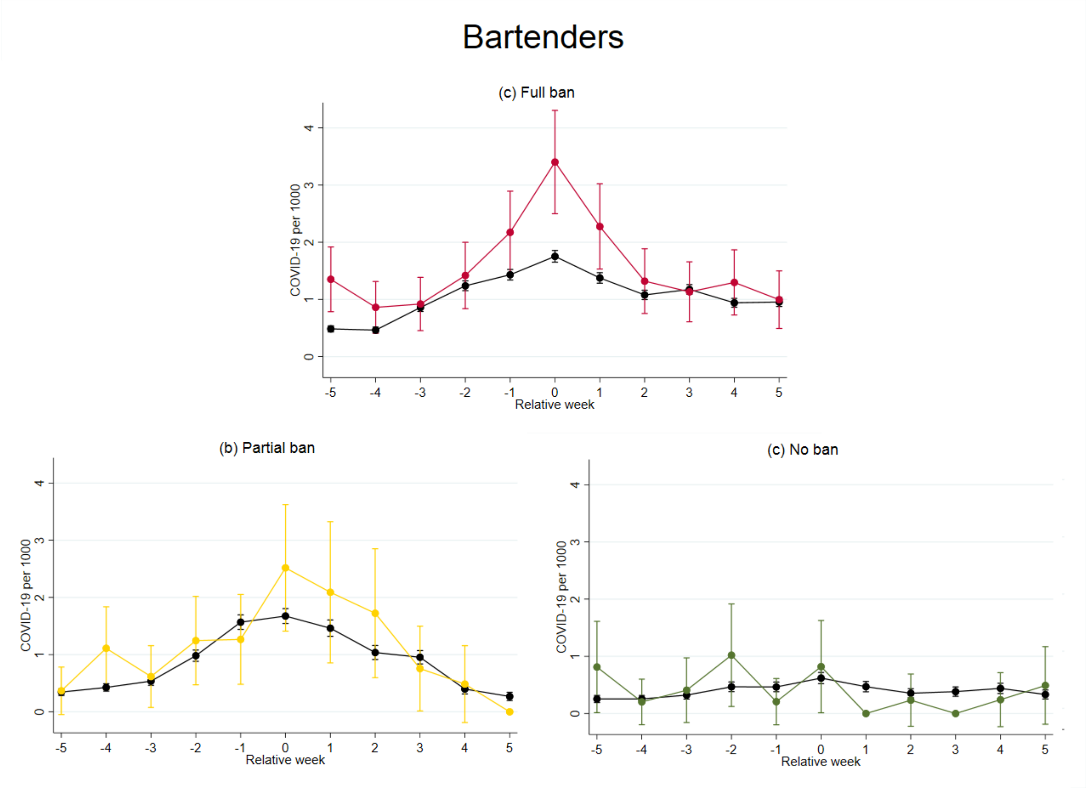


**Supplementary Figure B:** Estimated rates of confirmed COVID-19 for waiters separately (colored lines) and all other occupation (black lines) per week.

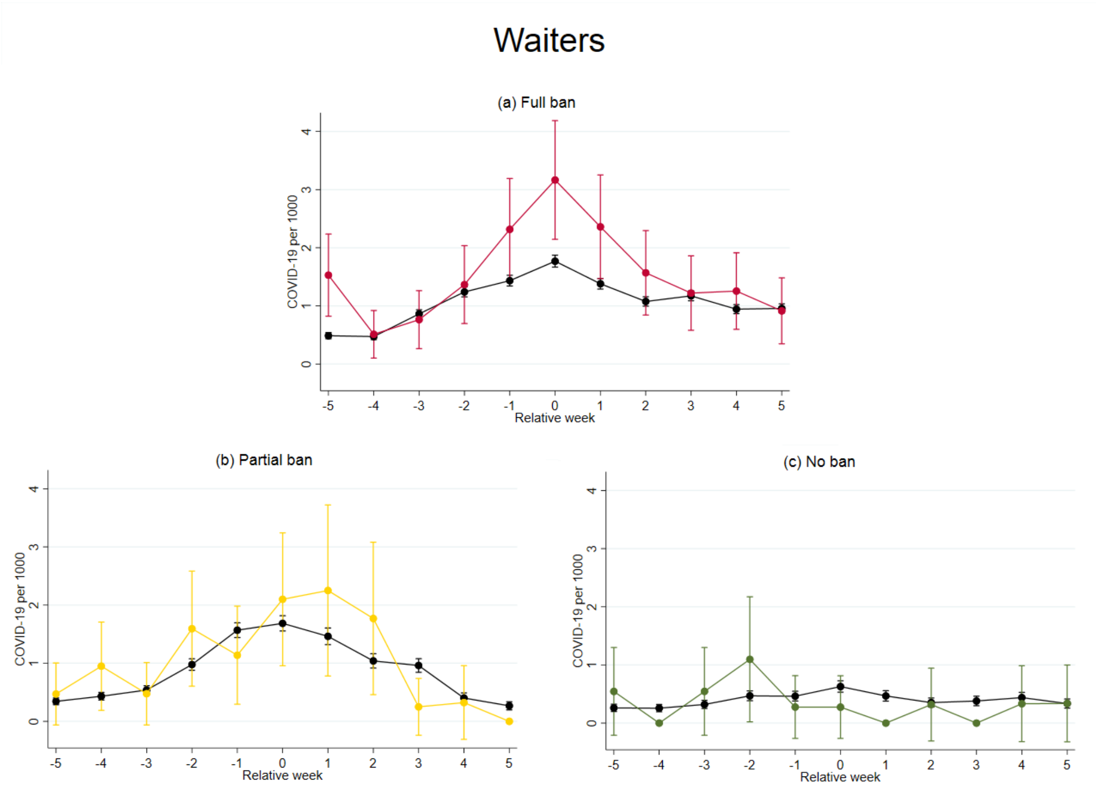
